# Using Google Health Trends to investigate COVID-19 incidence in Africa

**DOI:** 10.1101/2021.03.26.21254369

**Authors:** Alexander Fulk, Daniel Romero-Alvarez, Qays Abu-Saymeh, Jarron M. Saint Onge, A. Townsend Peterson, Folashade B. Agusto

**Author notes:** Corresponding author: Daniel Romero-Alvarez. Biodiversity Institute and Department of Ecology & Evolutionary Biology, 1345 Jayhawk Blvd. Lawrence, Kansas, 66045. United States.

## Abstract

The COVID-19 pandemic has caused over 350 million cases and over five million deaths globally. From these numbers, over 10 million cases and over 200 thousand deaths have occurred on the African continent as of 22 January 2022. Prevention and surveillance remain the cornerstone of interventions to halt the further spread of COVID-19. Google Health Trends (GHT), a free Internet tool, may be valuable to help anticipate outbreaks, identify disease hotspots, or understand the patterns of disease surveillance.

We collected COVID-19 case and death incidence for 54 African countries and obtained averages for four, five-month study periods in 2020-2021. Average case and death incidences were calculated during these four time periods to measure disease severity. We used GHT to characterize COVID-19 incidence across Africa, collecting numbers of searches from GHT related to COVID-19 using four terms: ‘coronavirus’, ‘coronavirus symptoms’, ‘COVID19’, and ‘pandemic’. The terms were related to weekly COVID-19 case incidences for the entire study period via multiple linear regression analysis and weighted linear regression analysis. We also assembled 72 predictors assessing Internet accessibility, demographics, economics, health, and others, for each country, to summarize potential mechanisms linking GHT searches and COVID-19 incidence.

COVID-19 burden in Africa increased steadily during the study period as in the rest of the world. Important increases for COVID-19 death incidence were observed for Seychelles and Tunisia over the study period. Our study demonstrated a weak correlation between GHT and COVID-19 incidence for most African countries. Several predictors were useful in explaining the pattern of GHT statistics and their relationship to COVID-19 including: log of average weekly cases, log of cumulative total deaths, and log of fixed total number of broadband subscriptions in a country. Apparently, GHT may best be used for surveillance of diseases that are diagnosed more consistently.

GHT-based surveillance for an ongoing epidemic might be useful in specific situations, such as when countries have significant levels of infection with low variability. Overall, GHT-based surveillance showed little applicability in the studied countries. Future studies might assess the algorithm in different epidemic contexts.

## INTRODUCTION

Coronavirus disease 2019 (COVID-19) is a respiratory disease caused by the severe acute respiratory syndrome coronavirus-2 (SARS-CoV-2) discovered in China in 2019. People infected experience a range of symptoms including headache, fever, difficulty breathing, and loss of taste and smell, or may be completely asymptomatic [1]. Since its discovery, SARS-CoV-2 has spread around the globe, with over 350 million confirmed cases as of 22 January 2022, according to John Hopkins University [2, 3]. The elderly (>65 years old), as well as those with pre-existing comorbidities, have the highest risk of mortality if infected [4]. COVID-19 spreads via respiratory particles, which allows it to infect others via contaminated aerosols and droplets suspended in the air in closed spaces [5]. Asymptomatic carriers account for a significant amount of secondary transmissions, with some reports showing that ~80% of infections may occur without symptoms, constituting the source of most secondary COVID-19 cases [1, 6, 7].

After the large-scale Ebola outbreak in 2015, African leaders were aware that swift and decisive action was needed to avert broad spread of COVID-19 and prevent healthcare system collapse. This awareness led to wide adoption of mitigation and control efforts that circumvented an overwhelming first epidemic wave with a partially structured continental response [8]. Regardless, testing in Africa has been limited: about 75% of COVID-19 diagnoses came from tests conducted in only 10 countries [5, 8]. The emergence of new of SARS-CoV-2 variants (e.g., Beta, Delta, Omicron, etc.) has made it difficult to predict wave dynamics across the continent, echoing other regional contexts [9]. Finally, although vaccination campaigns have been promoted as the definitive solution to the pandemic [10], several countries have struggled with vaccine uptake [11]. Africa as a continent has received only about 6% of the roughly 9 billion doses manufactured so far, even though about 17% of the world’s population lives there [12]. Further, uptake has been limited, as only 10% of Africans have been fully vaccinated [13, 14]. Factors determining this context, include disinformation via social media [15]; lack of syringes; lack of health workers to administer vaccines, especially in rural areas [13]; limited government planning; limited testing; and halting efforts at vaccine allocation [16].

Given difficulties in obtaining accurate and timely data on case counts and other epidemiological metrics for COVID-19 worldwide [17, 18], the current pandemic represents an opportunity to use digital epidemiology tools to fill gaps in information. Infodemiology is an area of epidemiology that uses digital data to gain insight into disease dynamics [19, 20]. The digital data used for this method of surveillance vary widely and may or may have not been intended for epidemiological purposes, coming from unexpected sources such as restaurant receipts, Facebook posts, or Google search queries [21, 22, 23].

Google developed two specific algorithms to address infectious diseases, Google Flu Trends (GFT) in 2009 and Google Dengue Trends (GDT) in 2011 [21], which, after inquiries into their usefulness, were shut down in 2015 [24]. Currently, Google maintains two portals by which to harvest search query data: Google Trends (GT) and Google Health Trends (GHT). GT inquiries yield a ranked score from 0 to 100 based on the highest frequency of searches for a term in a particular time period. GHT provides search counts from a relative proportion of a random sample of the overall Google search dataset for any particular term in a selected time interval [25]. Both of these portals have limitations, such as possibly excluding certain groups (e.g., the elderly, rural residents, low income populations), lack of detail on who is searching certain terms, and no insight into the underlying motivations of the searches [26].

Digital tools have been used in many instances to predict disease incidence [27, 28, 29] including COVID-19. Kurian et al. (2020) evaluated the applicability of GT in predicting COVID-19 cases in the United States (U.S.) in a state-by-state analysis [30]. They found that certain keywords had a strong correlation with COVID-19 cases, and concluded that GT may be a useful tool for predicting COVID-19 outbreaks. Brodeur et al. (2021) used GT to see how lockdowns affected well-being in the U.S. [31]. Once lockdowns were implemented, well-being likely decreased, as searches for certain terms such as ‘stress,’ ‘suicide,’ and ‘worry’ increased over the lockdown period. Ahmad et al. (2020) used gastrointestinal-related symptom search terms to determine whether GT could predict COVID-19 incidence, and found correlations between the search terms and increases of COVID-19 cases in multiple regions across the U.S. with a four-week lag [32].

Here, we explored whether GHT search query data correlate with COVID-19 incidence at the country level in Africa, as a potential complementary source for more customary forms of COVID-19 surveillance. We decided to use GHT instead of GT given the semi-quantitative nature of the information recovered by GHT. We collected case and death data for 54 African countries, and used four COVID-19-related search terms (see below) for each country. We then assessed whether Internet access, demography, economic information, or health variables, could explain GHT usefulness. Lastly, we calculated a standardized volatility index to illuminate whether variability in the signal of case incidence led to less accurate predictions by GHT.

## METHODS

### COVID-19 incidence data

Daily COVID-19 new cases and death counts were obtained for all 54 African countries from 2 February 2020 to 25 September 2021. Country-level case data were obtained via the Johns Hopkins COVID-19 global time series on the pandemic [33]; data were constrained to lab-confirmed cases only. We explored the progression of average daily COVID-19 case and death incidence per 100,000 people in Africa in four time periods, each roughly five months (~150 days) long: (a) 2 February 2020 to 30 June, (b) 1 July to 30 November, (c) 1 December to 30 April 2021, and (d) 1 May to 25 September 2021. We then converted daily new cases into weekly new cases for each of the countries to match the weekly GHT data up to 25 September 2021, for a total of 86 observations. We calculated weekly incidence rates by dividing the number of cases per week by the total population per country in millions [34]. Country-level population data were collected from the forecasted midyear 2020 estimates from the U.S. Census Bureau [35].

### Google Health Trends data

We downloaded data corresponding to four English terms from the GHT application programming interface (API): ‘coronavirus,’ ‘coronavirus symptoms,’ ‘COVID19,’ and ‘pandemic’. Although the four terms are related conceptually, they have the potential to capture a broad spectrum of information related to the disease [25, 36]. A simple and specific model is required here to maximize the usefulness of limited GHT data, which is why we have chosen only four terms closely related to the disease to fit the models. We addressed potential language barriers by collecting data for the latter two terms in French and Portuguese. The former two search terms were spelled the same in French and Portuguese, aside from accents, so the English versions of those terms captured a majority of individuals searching those terms in those other languages. We matched the relative search proportions of these words—which is the raw output provided by GHT [25]—with the weekly COVID-19 case incidence for the selected time period.

### Statistical analysis

We used a multiple linear regression model fitted with the four GHT English search terms as predictors of COVID-19 incidence at the country level for each of the 54 African countries being evaluated. We then performed the same analysis, substituting the latter two terms for their equivalents in French or Portuguese if a country had one of these listed as an official or spoken language by Nations Online [37]. The primary outcome measure was the adjusted *R*^*2*^ statistic, and we collected whichever adjusted *R*^*2*^ value was larger (in absolute value) from the models with all English or English and French/Portuguese terms. If one or more of the four terms chosen did not retrieve search counts from GHT, it was removed from the analysis for that country. At least two terms were included for each region. Multicollinearity may exist in our time series, but it will not affect prediction capabilities or goodness-of-fit [38]. Finally, to address possible autocorrelation and heteroskedasticity issues in our time series, we performed first-order differencing and ran the analyses again with a weighted least squares regression model, giving larger weight to those observations with lower variance. We collected the results from this weighted regression for a more conservative measure.

Next, we used the adjusted *R*^*2*^ statistics collected from the 54 African countries as our dependent variable and explored whether different categories of predictors might explain the pattern obtained. This analysis was conducted for the adjusted *R*^*2*^ statistics collected from the basic fitted regression models and the weighted regression models separately. Predictors for the African countries included Internet access, demographic, economic, and health indicators (Table 1); data were gathered from the World Bank [39]. We explored logarithmic transformations of each of these predictors to determine whether normalization of the indicators led to stronger correlations. Finally, we included as a predictor a standardized volatility index calculated using the standardized normalized case incidence data of each country as follows:

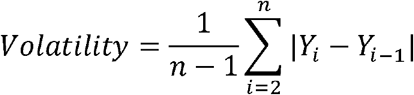

in which *n* is the total number of observations and *Y* is the normalized case incidence per country. The average of the absolute difference (i.e., volatility) summarizes the case signal reflecting if it is relatively constant or fluctuates broadly from week to week [25]. Overall, we explored a total of 72 potential explanatory variables (Table 1 and Supplementary Table 1).

**Table 1.**
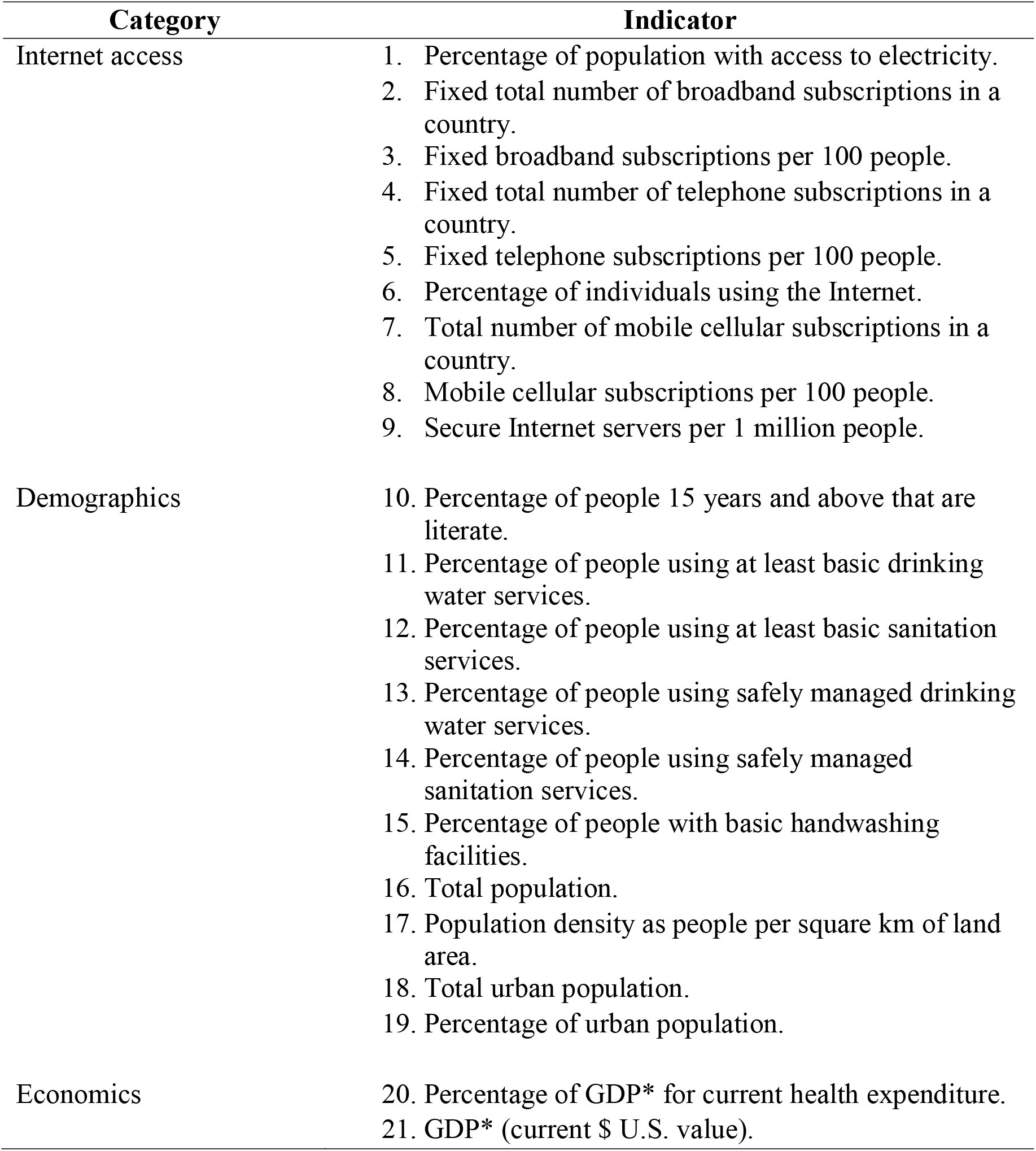
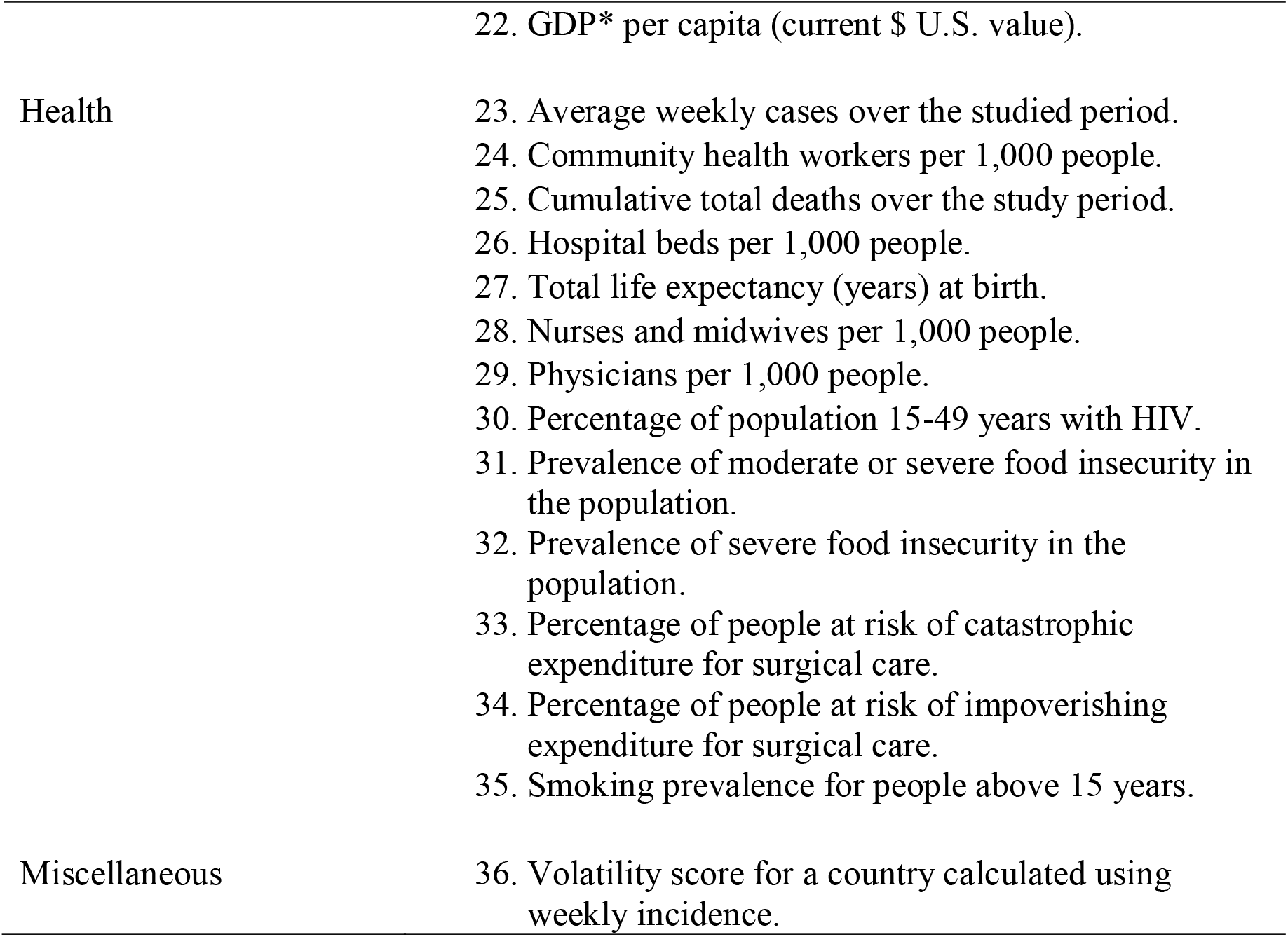
Predictors explored in the present study. Different categories were selected based on their perceived potential to explain patterns of Google Health Trends and COVID-19 regression models. We also evaluated the log of each predictor, giving for a total of 72 variables. *GDP = gross domestic product; HIV = human immunodeficiency virus. Raw values of the variables can be found in Supplementary Table 1.

| Category | Indicator |
| --- | --- |
| Internet access | 1. Percentage of population with access to electricity.<br>2. Fixed total number of broadband subscriptions in a country.<br>3. Fixed broadband subscriptions per 100 people.<br>4. Fixed total number of telephone subscriptions in a country.<br>5. Fixed telephone subscriptions per 100 people.<br>6. Percentage of individuals using the Internet.<br>7. Total number of mobile cellular subscriptions in a country.<br>8. Mobile cellular subscriptions per 100 people.<br>9. Secure Internet servers per 1 million people. |
| Demographics | 10. Percentage of people 15 years and above that are literate.<br>11. Percentage of people using at least basic drinking water services.<br>12. Percentage of people using at least basic sanitation services.<br>13. Percentage of people using safely managed drinking water services.<br>14. Percentage of people using safely managed sanitation services.<br>15. Percentage of people with basic handwashing facilities.<br>16. Total population.<br>17. Population density as people per square km of land area.<br>18. Total urban population.<br>19. Percentage of urban population. |
| Economics | 20. Percentage of GDP* for current health expenditure.<br>21. GDP* (current \$ U.S. value). |
| | 22. GDP* per capita (current \$ U.S. value). |
| Health | 23. Average weekly cases over the studied period.<br>24. Community health workers per 1,000 people.<br>25. Cumulative total deaths over the study period.<br>26. Hospital beds per 1,000 people.<br>27. Total life expectancy (years) at birth.<br>28. Nurses and midwives per 1,000 people.<br>29. Physicians per 1,000 people.<br>30. Percentage of population 15-49 years with HIV.<br>31. Prevalence of moderate or severe food insecurity in the population.<br>32. Prevalence of severe food insecurity in the population.<br>33. Percentage of people at risk of catastrophic expenditure for surgical care.<br>34. Percentage of people at risk of impoverishing expenditure for surgical care.<br>35. Smoking prevalence for people above 15 years. |
| Miscellaneous | 36. Volatility score for a country calculated using weekly incidence. |

Variables were analyzed individually using a pair-wise univariate linear regression and collectively in a stepwise regression, in which predictors were added and removed iteratively to obtain a subset of predictors that provided the best model outcome according to the Akaike Information Criterion (AIC). In addition, variables were analyzed using a least absolute shrinkage and selection operator (i.e., LASSO) regression for both untransformed and log-adjusted data to avoid overfitting and produce simpler models. Countries with missing variable information were removed from the univariate regression including that particular variable (38/72; 53% of variables had at least one country removed, Supplementary Table 1), and only variables with information for every country were used in the stepwise and LASSO regressions. All analyses were done for both adjusted *R*^*2*^ values collected from the basic regression and the weighted regression. All analyses were performed in R [40]. Data and scripts to replicate the results of this study are available in a GitHub repository accompanying this publication (https://github.com/alxjfulk/GHT-and-COVID19-code).

## RESULTS

Examining the distribution of first cases among the 54 African countries, we observed that dates of first reported COVID-19 cases were centered around March 2020. Egypt (EGY) reported the first case of COVID-19 on the continent on 14 February 2020, 15 days after the World Health Organization (WHO) declared the COVID-19 epidemic an emergency of international concern [41]. Comoros (COM) and Lesotho (LSO) were the last countries to report COVID-19 introductions, with first cases on 30 April and 13 May 2020, respectively (Figure 1).

**Figure 1.**
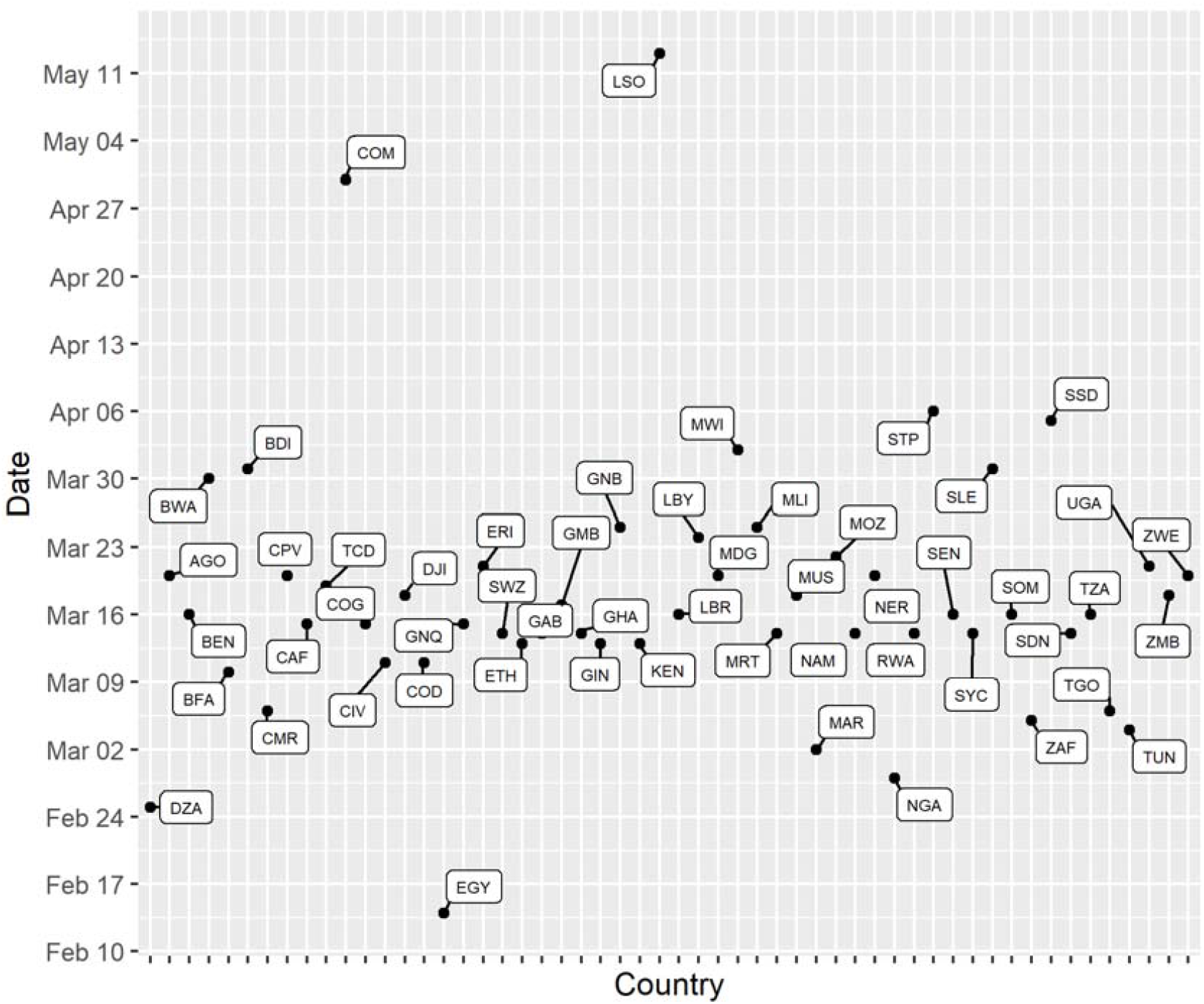
Distribution of the day of the first COVID-19 reported case in 54 African countries. The plot depicts the dates of the first reports of COVID-19 cases in the 54 studied African countries as reported by the Johns Hopkins global time series on the pandemic (CRC, 2020; Dong et al, 2020). The countries in this distribution are designated by their three-letter Alpha-3 codes: DZA: Algeria, AGO: Angola, BEN: Benin, BWA: Botswana, BFA: Burkina Faso, BDI: Burundi, CPV: Cabo Verde, CMR: Cameroon, CAF: Central African Republic, TCD: Chad, COM: Comoros, COD: Democratic Republic of the Congo, COG: Congo, CIV: Côte d’Ivoire, DJI: Djibouti, EGY: Egypt, GNQ: Equatorial Guinea, ERI: Eritrea, SWZ: Eswatini, ETH: Ethiopia, GAB: Gabon, GMB: Gambia, GHA: Ghana, GIN: Guinea, GNB: Guinea-Bissau, KEN: Kenya, LSO: Lesotho, LBR: Liberia, LBY: Libya, MDG: Madagascar, MWI: Malawi, MLI: Mali, MRT: Mauritania, MUS: Mauritius, MAR: Morocco, MOZ: Mozambique, NAM: Namibia, NER: Niger, NGA: Nigeria, RWA: Rwanda, STP: Sao Tome and Principe, SEN: Senegal, SYC: Seychelles, SLE: Sierra Leone, SOM: Somalia, ZAF: South Africa, SSD: South Sudan, SDN: Sudan, TZA: United Republic of Tanzania, TGO: Togo, TUN: Tunisia, UGA: Uganda, ZMB: Zambia, ZWE: Zimbabwe.

Countries with highest COVID-19 case incidences for the first time period include Djibouti (3.39 cases per 100,000 people), São Tome and Principe (2.25), and South Africa (1.79) (Fig. 2). During the second period, Cameroon (10.7), Libya (7.78), and South Africa (7.39) were most affected (Fig. 2). For the third and fourth periods, countries across the continent reported increased COVID-19 incidences, with Seychelles (third period = 39.1; fourth period = 109), Tunisia (third period = 12.0; fourth period = 22.8), Botswana (third period = 10.3; fourth period = 37.8), Namibia (third period = 8.55; fourth period = 20.3), and South Africa (third period = 9.28; fourth period = 15.7) ranking top among the countries studied (Fig. 2). Tanzania had an incidence of 0 for the second and third time periods, which will be discussed below.

**Figure 2.**
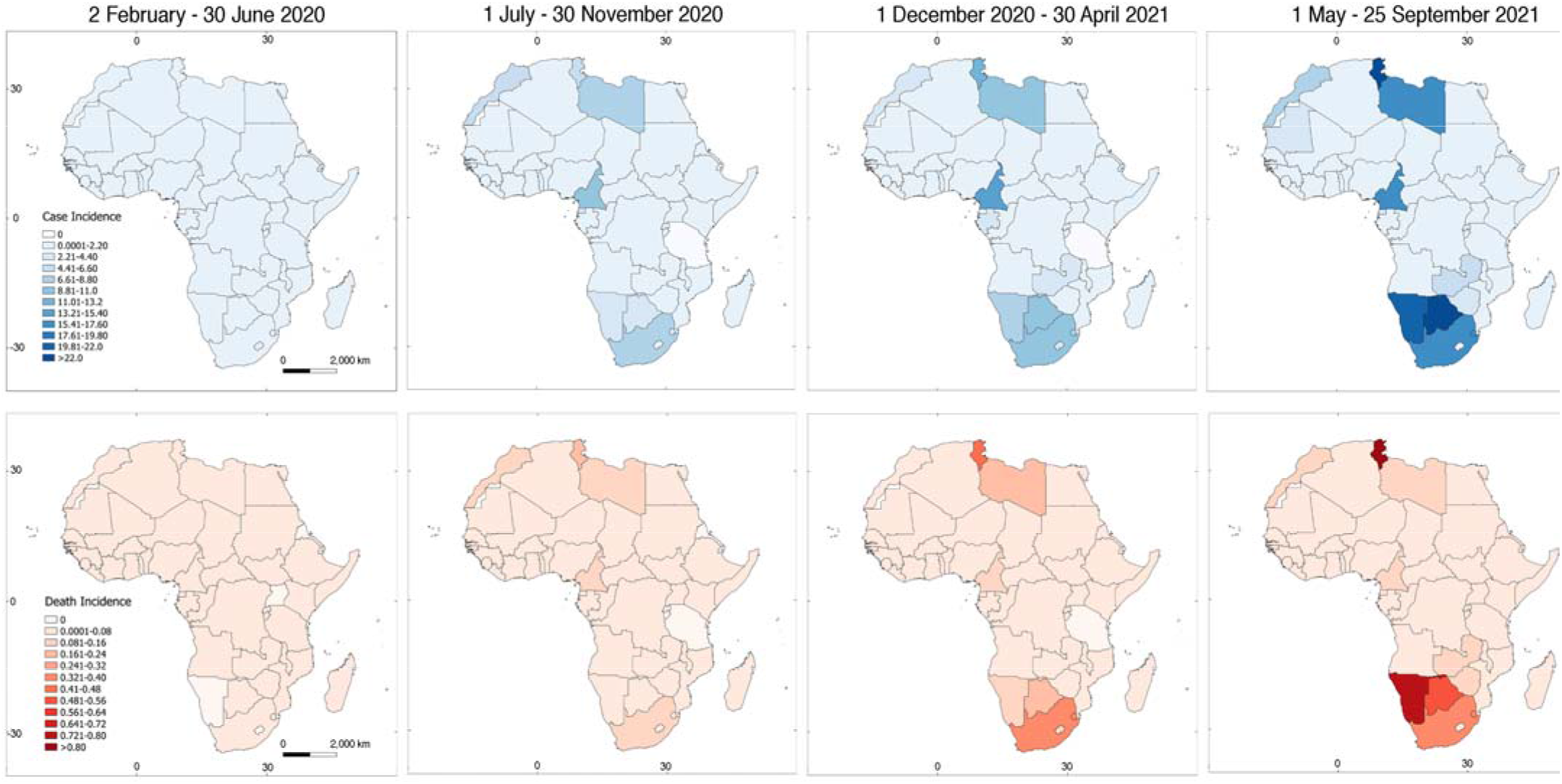
Average case and death incidences of COVID-19 per 100,000 people over four five-month time periods in Africa. Eight plots show average case incidences (upper panels) and average death incidences (bottom panels) over four five-month time periods from 2 February 2020 to 25 September 2021. Scale is the same for all case/death incidence maps and is depicted in the left panels; numbers are individuals affected per 100,000 people.

COVID-19 death incidence was recorded for all the African countries in the second time period except for Eritrea, Seychelles, Comoros, Mauritius, Tanzania, and Burundi, although the latter four reported 5.51×10^−3^, 4.83×10^−3^, 2.39×10^−4^, and 5.62×10^−5^ death incidences per 100,000 people during the first period, respectively. Further, South Africa (0.219 deaths per 100,000 people) and Tunisia (0.179) were the countries reporting the highest death incidence in the second period. For the third period, highest death incidences were again reported in South Africa (0.385) and Tunisia (0.422); for the fourth period, highest incidences were recorded in Tunisia (0.808), Namibia (0.732), and Seychelles (0.584).

Few countries had no information for one or two of the chosen English terms (6/54; 11.1%); only ‘coronavirus’ and ‘COVID19’ always recovered search query counts. Several countries that had French or Portuguese listed as an official language returned no information for either one or both language-specific terms (8/32; 25%, Supplementary Table 2). Overall, the adjusted *R*^*2*^ values collected to depict the relationship between GHT search queries and COVID-19 weekly incidence were low, never above 0.4 for any of the countries in either the basic regression or the weighted regression (Fig 3). The largest adjusted *R*^*2*^ results from the basic regression were for Algeria (0.33), Ethiopia (0.20), and Kenya (0.19; Fig. 4). The countries with the lowest adjusted *R*^*2*^ results from the basic regression included Burkina Faso (−0.028), Sierra Leone (−0.030), and Sudan (−0.031; Figs. 3, 4, and Supplementary Table 2). For the weighted regression analysis on the first-order differenced case incidence and GHT data, the countries that returned the largest adjusted *R*^*2*^ results were Guinea-Bissau (0.24), Lesotho (0.08), and Niger (0.07, Fig. 3), respectively. The lowest adjusted *R*^*2*^ results came from Zimbabwe, Egypt, and Mauritania each with an adjusted *R*^*2*^ value of −0.05 (rounded; see Supplementary Figures).

**Figure 3:**
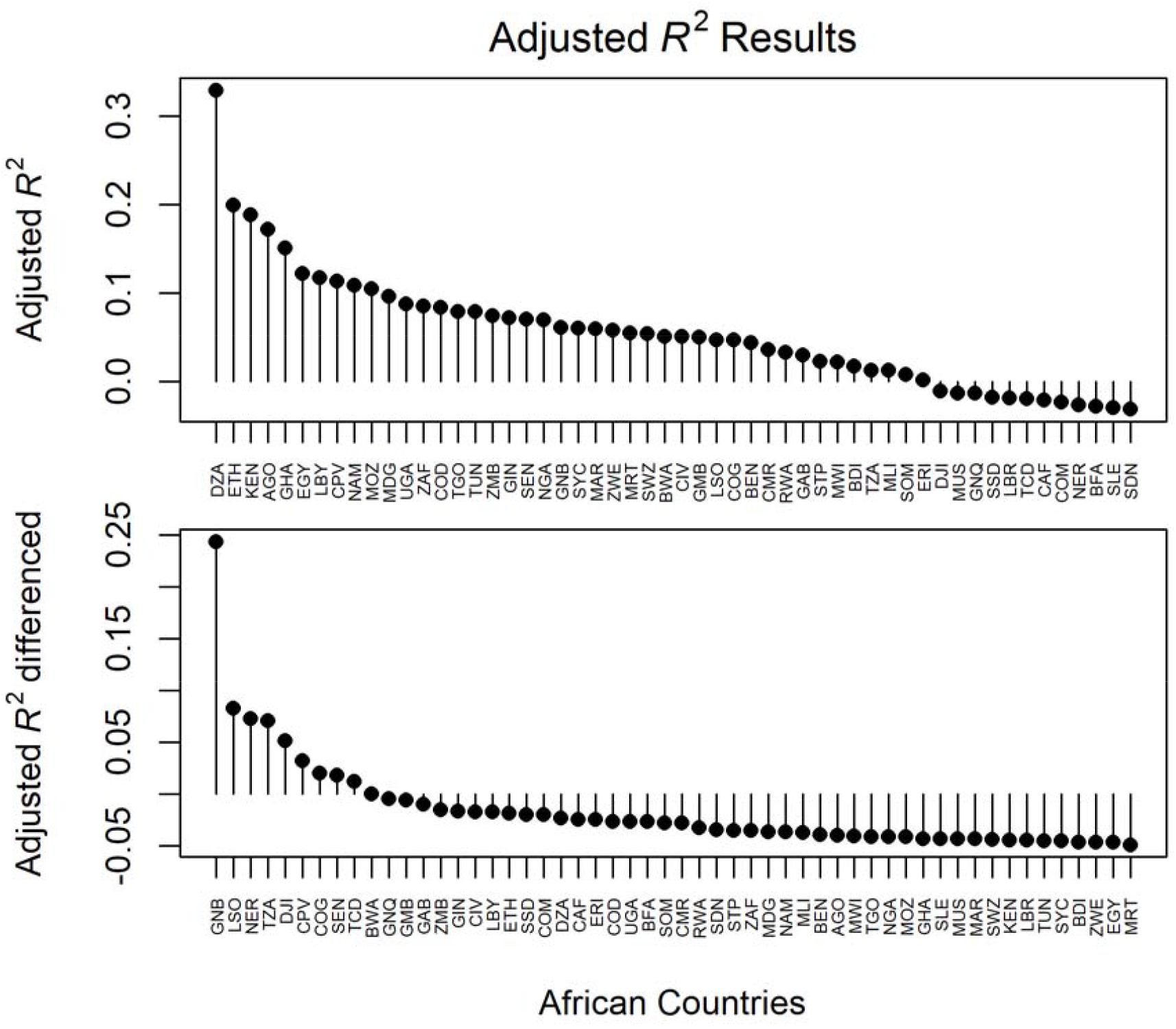
Results of multiple linear regression analysis between COVID-19 incidence and Google Health Trends (GHT) search terms. The adjusted *R*^*2*^ of the basic (upper panel) and the weighted (bottom panel) regression analysis is depicted here to visually represent the countries from the highest to lowest performance. The countries in this figure are designated by their three-letter Alpha-3 codes as in Figure 1.

**Figure 4.**
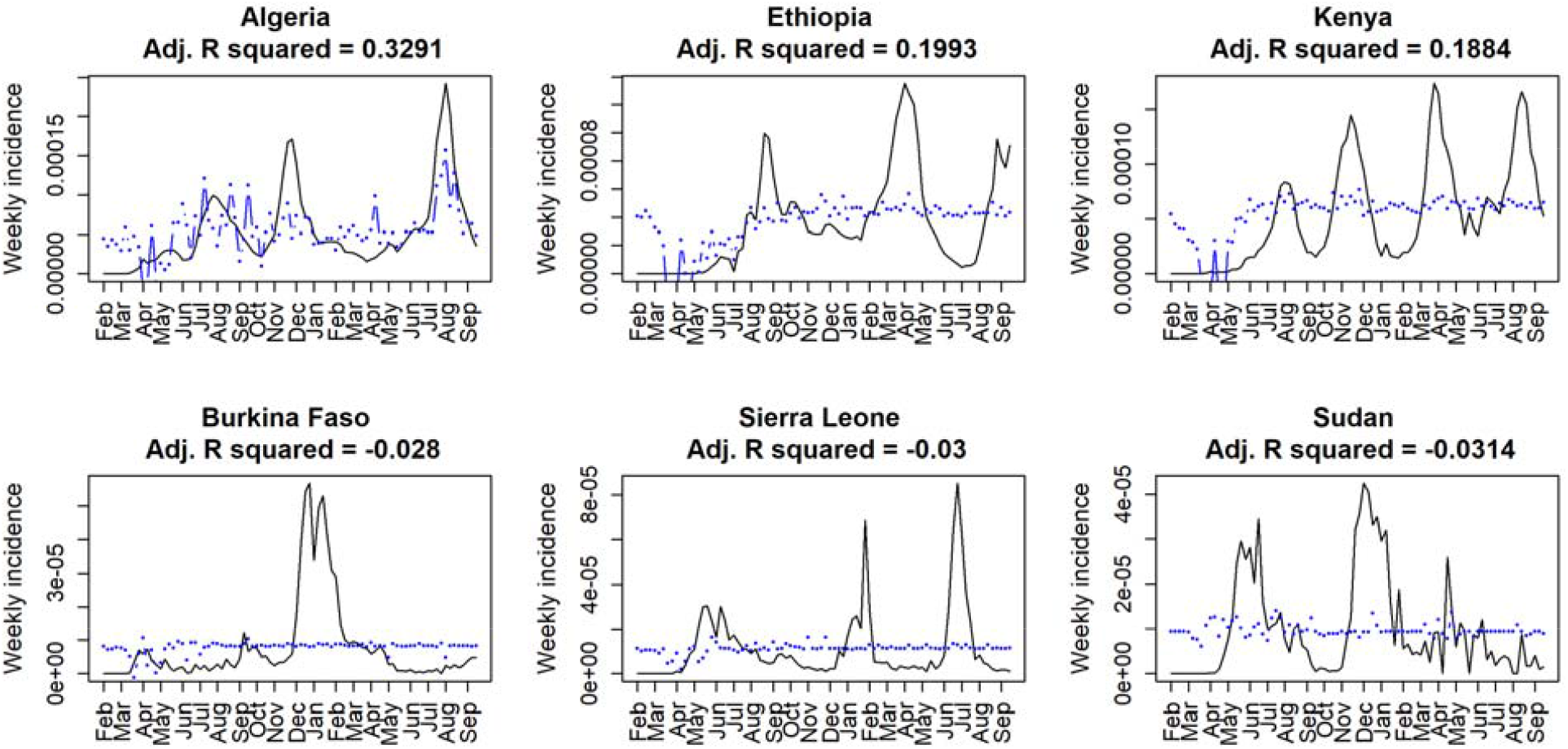
Best and worst performing countries from the basic regression analysis of Google Health Trends (GHT) data. When analyzing whether GHT correlated with case incidence (black line) via a multiple linear regression analysis (blue line), the three best performing countries were Algeria (DZA), Ethiopia (ETH), and Kenya (KEN), respectively (upper panels). The three worst performing countries were Burkina Faso (BFA), Sierra Leone (SLE), and Sudan (SDN), respectively (bottom panels).

Several of the 72 indicators were able to predict at least in part the pattern of adjusted *R*^*2*^ statistics obtained for the 54 African countries. Almost all univariate, linear analyses from the basic regression yielded adjusted *R*^*2*^ values of 0.25 or less, except for the log of average weekly cases (0.37), log of cumulative total deaths (0.30), and log of fixed total number of broadband subscriptions in a country (0.26, Supplementary Table 3). The only adjusted *R*^*2*^ value greater than 0.25 from the weighted regression analyses came from the number of community health workers per 1,000 people, but that variable only had data for 26 countries. The log of average weekly cases, the log of GDP, and the log of the volatility scores were also statistically significant, though they yielded low adjusted *R*^*2*^ values (*R*^*2*^ = 0.120, 0.057, and 0.080, respectively; *p* = 0.0059, 0.046, and 0.022, respectively). The stepwise regression analysis on the untransformed data showed that a model including percentage of GDP for current health expenditure, life expectancy (years) at birth, mobile cellular subscriptions per 100 people, total population, GDP per capita, percentage of people using the Internet, total urban population, total number of mobile cellular subscriptions in a country, average weekly cases over the studied period, and (notably) volatility score for a country calculated using weekly incidence yielded an adjusted *R*^*2*^ of 0.40. LASSO regression analysis revealed that a model with life expectancy (years) at birth showed an adjusted *R*^*2*^ of 0.13.

When going further with the analysis using the adjusted *R*^*2*^ values collected from the weighted regression model, the volatility score for a country calculated using weekly incidence was deemed most useful for both stepwise and LASSO regressions, though the latter method also returned percentage of the population with access to electricity (*R*^*2*^ = 0.051, and 0.063, respectively). Conversely, using logarithmically transformed variables, a stepwise regression model including average weekly cases over the studied period, percentage of GDP for current health expenditure, life expectancy (years) at birth, yielded an adjusted *R*^*2*^ value of 0.47. Using the adjusted *R*^*2*^ values collected from the weighted regression analyses, a model including average weekly cases over the time period studied was selected using the stepwise regression. LASSO regression analysis of logarithmic transformed variables indicated that a model including percentage of individuals with access to electricity, life expectancy (years), average weekly cases over the period studied, cumulative deaths, percentage of GDP for current healthcare expenditure, and total population gave the highest adjusted *R*^*2*^ of 0.45. Using the adjusted *R*^*2*^ collected from the weighted regression analyses, volatility score for a country calculated using weekly incidence and average weekly cases were returned (*R*^*2*^ = 0.13). The results of these models yielded adjusted *R*^*2*^ values larger than most of the univariate analyses (Supplementary Table 3); nevertheless, we are cautious in our interpretation of these results [25, 42, 43].

## DISCUSSION

Despite successful demonstrations of the GHT algorithm to aid infectious disease surveillance for influenza, dengue, and other diseases [26, 43, 44], our study demonstrates that, in the context of the COVID-19 epidemic, GHT appeared to be difficult to implement as a predictor of COVID-19 incidence and impact. Average weekly cases over the period studied was an important predictor when analyzing possible patterns in the adjusted *R*^*2*^ values collected from both the basic regression and weighted regression analyses. The volatility score for a country was also an important predictor of the applicability of GHT, as demonstrated in our univariate, stepwise, and LASSO models (Supplementary Table 3). Finally, indicators related to Internet access (mobile cellular subscriptions per 100 people, total number of mobile cellular subscriptions in a country, percentage of individuals using the Internet, percentage of individuals with access to electricity), health (life expectancy (years) at birth), demographics (total population, total urban population), and economics (percentage of GDP for current health expenditure, GDP, GDP per capita) were selected heterogeneously with different modeling approaches (Supplementary table 3).

The top three ranking countries based on adjusted *R*^*2*^ values in the basic regression (Algeria, Ethiopia, and Kenya) all seemed to have similar COVID-19 incidence signal type (Fig. 4, upper panels). Cases begin at zero, spike, and subsequently drop to a lower, but still significant level of incidence, followed by additional waves, potentially reflecting an exhaustion of susceptible individuals or dynamics of new variants [45, 46]. Algeria, Ethiopia, and Kenya all had strong responses to initial outbreaks of COVID-19 and invested significantly in preventative measures against COVID-19 such as testing, vaccination, and healthcare [47, 48, 49]. These three countries also ranked within the top 10 when looking at the total number of mobile cellular subscriptions in a country and GDP (Supplementary table 1). On the other hand, Burkina Faso, Sierra Leone, and Sudan are lower-income countries, and have struggled to combat COVID-19 [50, 51, 52]; according to World Bank data, they ranked lower than the top-ranking countries in terms of total number of mobile cellular subscriptions in a country (Supplementary table 1). Furthermore, four out of these six countries had an extremely low percentage of individuals using the Internet (< 20% as of 2017), which may indicate that the way Internet access is currently measured reflects GHT behavior poorly. Interestingly, the three countries with the worst GHT prediction (Fig. 4, lower panels) showed fewer cases and greater variability in their incidence signal compared to the best-performing countries. The combination of these results may indicate that some consistent level of infection is required for keeping the interest of communities searching information through Google search engines. This gives GHT a chance to match cases, and it may perform better when a rapid growth of infection coincides with interest in the topic and Internet search volume for disease-specific terms is likely to be high, regardless of the level of penetration of Internet access. Finally, GHT seemed to perform best in those countries that have infrastructure to support accurate case counts.

As in the rest of the world, incidence of both COVID-19 cases and COVID-19 related deaths increased across Africa steadily during the study period. However, in the second and third periods of our study, Tanzania showed zero COVID-19 cases (Fig. 2). Upon closer examination, the country stopped reporting coronavirus cases and deaths in April of 2020, so any patterns that might be observed for Tanzania (0.012) are actually reflecting a lack of data [53]. While COVID-19 numbers are concerning on the continent, Africa has been observed to have a lower disease burden in comparison to other regions of the world [54–57, but see 58]. As of August 2020, Africa had reported approximately 69.2 cases and 1.31 deaths per 100,000 people in nearly seven months since COVID-19 was declared a pandemic; For comparison, the U.S. at that point in time had seen roughly 1500 cases per 100,000 people, and Brazil confirmed roughly 47.0 deaths per 100,000 people [2, 3].

Although infodemiology approaches represent the next frontier of infectious disease surveillance [19, 23], the present modeling effort demonstrates that search queries from GHT are difficult to correlate with incidence of disease in the context of an emerging epidemic. In contrast with diseases such as influenza or dengue that are studied consistently in a seasonal pattern or are endemic to multiple regions [25, 43, 59], COVID-19 represented an unprecedented case study that might render Google-based information mining ineffective for several reasons: (a) partial or incomplete COVID-19 case detection and reporting [8, 60], (b) media-induced search behavior [61], or even (c) information fatigue [36]. Thus, we encourage caution regarding interpretation of COVID-19 modeling experiments based on Google search engines. For example, Ahmad et al. (2020) found a correlation between gastrointestinal search terms obtained through GT and COVID-19 cases and suggested that Internet searches may be useful in predicting COVID-19 cases using a four-week lag in the U.S. [32]. This correlation, however, might be an artifact since none of the gastrointestinal terms is specific to COVID-19, and the only COVID-19 specific term—’ageusia’—increased during the time that the pandemic was declared (i.e., 11 March) and decreased while cases started to increase (Figure 1 in [32]). The U.S. showed an increase in case numbers driven by increasing test capacity, thus, these case numbers were reflecting disease incidence inaccurately [62]. Thus, although our findings are based on the GHT algorithm, we are cautious about interpreting our results and those of others in characterizing COVID-19 via Google search engines. Similar to our findings, Asseo et al. (2020) found correlations between GT search queries related to smell and taste at the beginning of the pandemic in Italy and the U.S., which faded in succeeding epidemiological weeks [36]. More importantly, Asseo et al. (2020) also showed how correlation patterns fall apart when analyzing Google search queries and COVID-19 incidence in nonconsecutive weeks (e.g. 11-17 March vs. 1-7 April 2020 in [36]).

Some limitations of the present research are as follows. Because of the timeframe of the study and the availability of GHT data as weekly counts, we had to convert daily cases to weekly cases, limiting our analysis to only 86 observations, decreasing the statistical power of our approach. Moreover, the four terms related with COVID-19 that were selected might not be as popular in the region as expected. Language might be an important although permeable barrier [25, 27]. Still, in the present study the addition of French and Portuguese translations of search terms did not yield significantly higher adjusted *R*^*2*^ values (Supplementary Table 2). Finally, we lacked complete data for some of the predictors (e.g., prevalence of severe food insecurity in the population; Supplementary Table 3) which halts interpretation of several of the indicators used; however, those that were available for all the countries proved useful here as in other research studies (e.g., total population, signal volatility, disease incidence, etc) [25, 43].

## CONCLUSIONS

Surveillance for an ongoing epidemic via GHT might be useful in specific situations in which accurate case counts can be retrieved and there is sustained level of disease incidence. Google instruments to recover population search counts—GT and GHT—are potentially powerful digital epidemiology tools that can lead to greater insight into disease dynamics, and should be studied and implemented depending on the particular context of an outbreak [25, 30, 63–66]. Future directions to examine GHT on COVID-19 research include expansion of the analysis to a larger dataset both in time and space. Other refinements can be implemented, combining other forms of digital data (e.g., Twitter, Wikipedia) to determine if addition of more information improves the predictive power of the model.

## Data Availability

Data is available as a supplementary material in the current submission.

## ACKNOWLEDGMENTS

DRA thanks the writing-original-group (WOG) for support during the development of this paper.

## SUPPLEMENTARY MATERIAL

**Supplementary Table 1**. Raw data for the predictors used to explore patterns of Google Health Trends search queries and COVID-19 incidence in 54 African countries.

**Supplementary Table 2**. Date of the first COVID-19 case reported in Africa. Results of multiple linear regression analysis performed between COVID-19 incidence and Google Health Trends search queries from four selected terms.

**Supplementary Table 3**. Univariate and multivariate linear regression analysis to explore associations between adjusted *R*^*2*^ of Google Health Trends search queries and COVID-19 incidence in 54 African countries.

**Supplementary Figures**. Plots depicting the best and worst performing countries in the weighted regression analysis and plots depicting the multiple linear regression models between COVID-19 case counts and Google Health Trends search queries for the 54 African countries studied in the present manuscript.

